# Association Between Minimum Temperature and Carbapenem-Resistant *Enterobacteriaceae* Infections in South Korea, 2018–2021: A Retrospective Time-Series Analysis

**DOI:** 10.1101/2025.09.10.25335271

**Authors:** Jeehyun Kim, Derek R MacFadden, Byung Chul Chun, Mauricio Santillana

## Abstract

Carbapenem-resistant Enterobacteriaceae (CRE) infections pose a major public health threat with limited treatment options and high mortality. Using national surveillance data from South Korea (2018–2021), we conducted a time-series analysis to assess associations between monthly CRE incidence, minimum temperature, and meropenem usage. A 10°C rise in minimum temperature was associated with a 9.4% increase in CRE incidence (95% Confidence Interval: 5.0–13.9%). Findings suggest temperature as an environmental driver of antimicrobial resistance, supporting integration of climate data into surveillance.

---

Carbapenem-resistant *Enterobacteriaceae* (CRE) infections pose a global threat due to limited treatment options and high mortality, as carbapenems are considered last-resort antibiotics for multidrug-resistant Gram-negative infections and CRE strains are frequently resistant to multiple different antibiotic classes [1,2].

While antibiotic use drives resistance, recent studies suggest that environmental factors, particularly temperature, may also influence antimicrobial resistance (AMR) across geographic regions [3–5]. However, high-resolution temporal studies capturing seasonal infection changes at specific locations are needed to clarify the role of temperature in AMR infections [3–5]. We investigate the association between monthly CRE incidence, ambient temperature, and meropenem usage using nationwide population-based data from South Korea.

## Data collection

We conducted a retrospective time-series study using national CRE surveillance data from January 2018 to December 2021. In South Korea, CRE is a notifiable infectious disease, requiring reporting of confirmed cases and colonization within 24 hours [1]. Monthly CRE infection rates per 100,000 population were calculated and log-transformed. Minimum land surface temperature (°C) was derived from MODIS satellite data and summarized monthly. Quarterly meropenem usage, expressed as days of therapy (DOT) per 1,000 inhabitants per day, was extracted from the 2022 Korea National Antimicrobial Use Analysis System report as measure of carbapenem use.

## CRE in South Korea, 2018–2021

A total of 68,747 CRE cases were reported with an increasing trend from 2018 to 2021 (Table 1). Most cases were male (39,058, 56.8%) and aged 70–79 years (19,252, 28.0%). CRE incidence peaked between July and September (Table 1).

**Table 1.** Annual CRE Case Counts and Monthly Case Distribution with Incidence per 100,000 Population in South Korea, 2018–2021.

|  | 2018 | 2019 | 2020 | 2021 |
| --- | --- | --- | --- | --- |
| Total cases | 11,954 | 15,369 | 18,113 | 23,311 |
| Incidence rate (per 100,000) | 23.2 | 29.7 | 34.9 | 45.1 |
| **Monthly case counts** |  |  |  |  |
| January | 883 | 1,178 | 1,455 | 1,707 |
| February | 744 | 929 | 1,306 | 1,415 |
| March | 859 | 970 | 1,159 | 1,626 |
| April | 901 | 1,086 | 1,231 | 1,708 |
| May | 915 | 1,171 | 1,337 | 1,697 |
| June | 1,005 | 1,123 | 1,623 | 2,052 |
| July | 1,141 | 1,521 | 1,853 | 2,381 |
| August | 1,293 | 1,714 | 1,723 | 2,510 |
| September | 1,115 | 1,464 | 1,703 | 2,307 |
| October | 1,159 | 1,637 | 1,680 | 2,175 |
| November | 975 | 1,309 | 1,569 | 1,901 |
| December | 964 | 1,267 | 1,474 | 1,832 |

## Time lag of meropenem usage

To determine the appropriate time lag of meropenem usage in relation to carbapenem-resistant *Enterobacteriaceae* (CRE) incidence, we conducted analyses using linear regression models. The dependent variable was the log-transformed incidence rate of CRE. Each model included an overall time trend (Date), meropenem usage with varying lags, and minimum temperature as covariates.

Three models were constructed using different lag structures for meropenem usage: no lag, a lag of one quarter (3 months), and a lag of two quarters (6 months). The model specification was as follows:

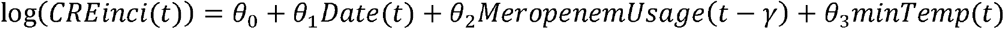

where γ = 0, 1, 2 represents the lag period in quarters.

The lag period was chosen according to the model that produced the largest coefficient for minimum temperature. This criterion identified the no-lag model (Table 2). Interestingly, the positive association remained significant across all models that considered multiple lags for meropenem use.

**Table 2.**
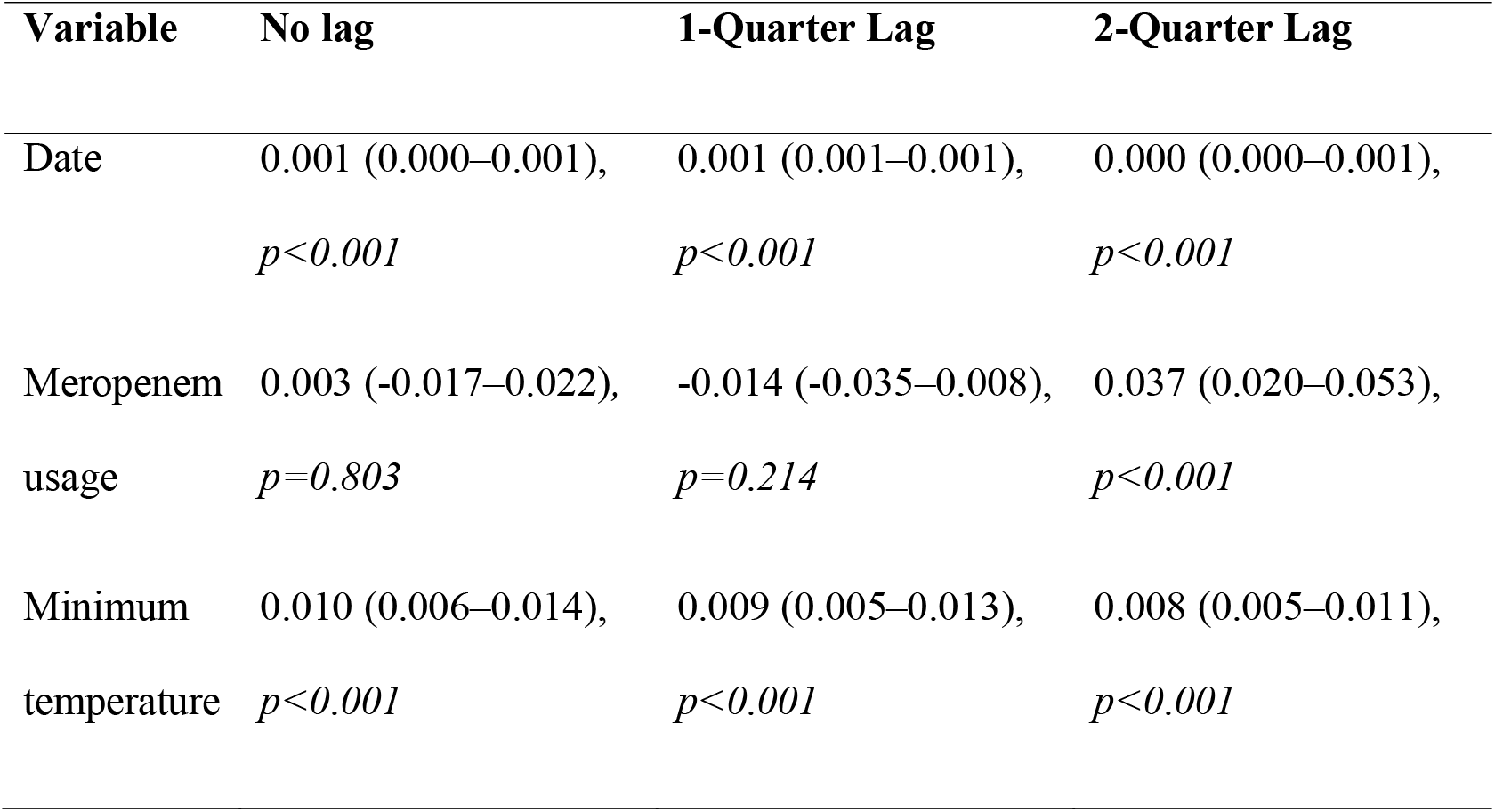
Sensitivity Analysis of Meropenem Usage Lag on Log-Transformed CRE Incidence, presented as coefficient (95% confidence interval), *p*-value.

| Variable | No lag | 1-Quarter Lag | 2-Quarter Lag |
| --- | --- | --- | --- |
| Date | 0.001 (0.000–0.001),<br>*p*<0.001 | 0.001 (0.001–0.001),<br>*p*<0.001 | 0.000 (0.000–0.001),<br>*p*<0.001 |
| Meropenem<br>usage | 0.003 (-0.017–0.022),<br>*p*=0.803 | -0.014 (-0.035–0.008),<br>*p*=0.214 | 0.037 (0.020–0.053),<br>*p*<0.001 |
| Minimum<br>temperature | 0.010 (0.006–0.014),<br>*p*<0.001 | 0.009 (0.005–0.013),<br>*p*<0.001 | 0.008 (0.005–0.011),<br>*p*<0.001 |

## Minimum temperature and CRE incidence

To evaluate the association between minimum temperature and CRE incidence, we used linear regression models adjusting for time trends and meropenem usage. We analyzed residuals against temperature:

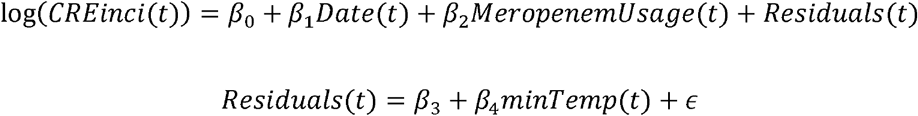

We found that a 10°C increase in minimum temperature was associated with a 9.4% increase in CRE incidence (95% Confidence Interval = 5.0–13.9%, *R*^2^=0.33, Figure 1).

**Figure 1.**
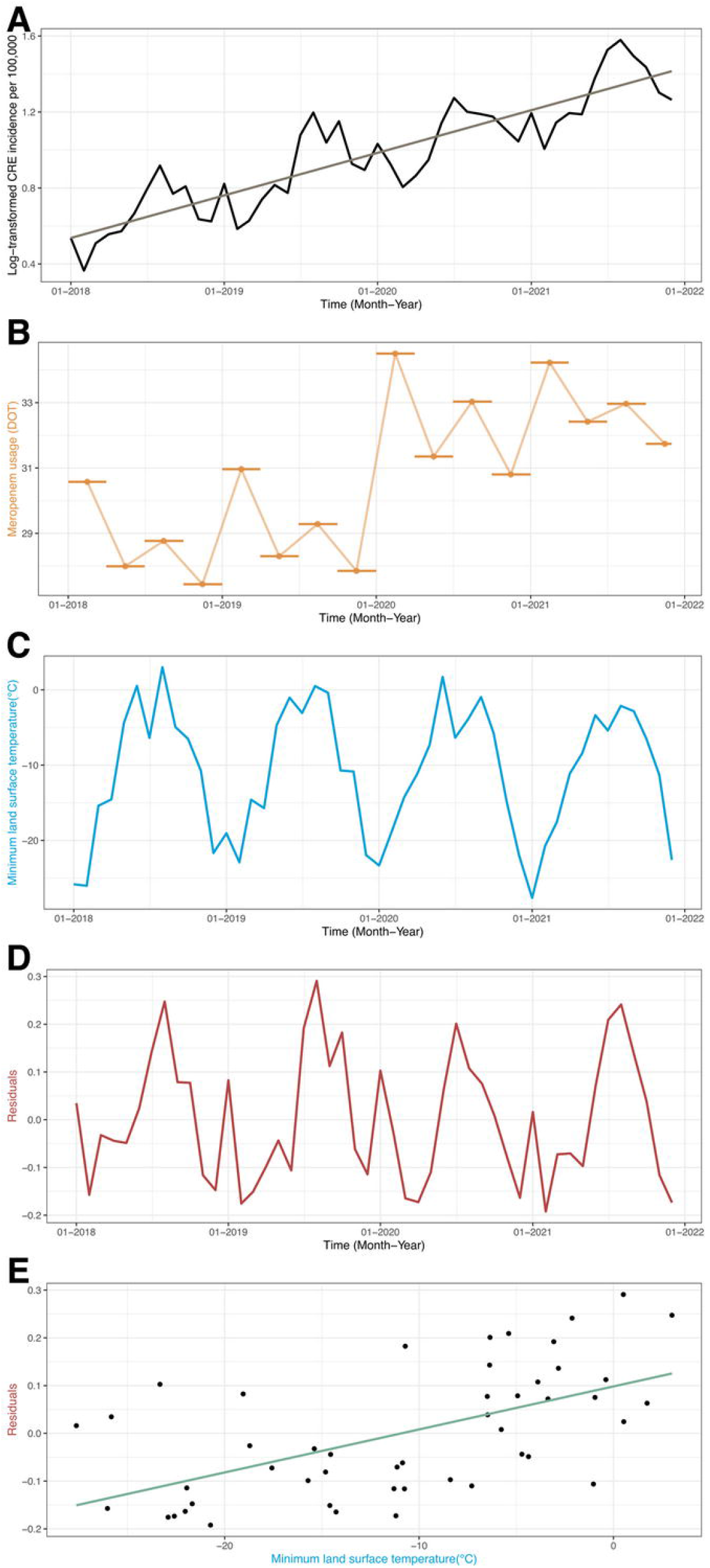
Time-Series Trends and Temperature Association with CRE Incidence in South Korea, 2018–2021. A. Log-transformed monthly CRE incidence per 100,000 population and its linear trend (β = 0.001, R^2^ = 0.79, *p <0.001*). B. Quarterly meropenem usage (days of therapy per 1,000 inhabitants per day). C. Monthly minimum land surface temperature (°C). D. Residuals from the linear model accounting for time trend and meropenem usage. E. Linear association between residuals of CRE incidence and minimum temperature, showing a positive relationship (β = 0.009, R^2^ = 0.33, *p <0.001*). CRE, carbapenem-resistant Enterobacteriaceae; DOT, days of therapy per 1,000 inhabitants per day.

## Discussion

Our findings demonstrate a positive association between ambient temperature and CRE incidence. The temperature effect was similar in scale to that previously reported for other classes of resistance [3], supporting the hypothesis that temperature may influence AMR within specific regions over time as well as across geographic locations.

By controlling for antibiotic usage, our findings build upon previous Korean hospital-based research reporting increased acquisition of carbapenemase-producing *Enterobacterales* during warm seasons [6]. Mechanisms are likely manifold, including microbial characteristics, seasonal behavior, and hospital water systems [2,6].

This study has limitations. We did not assess changes in carbapenem resistance prevalence among *Enterobacteriaceae*. Nonetheless, the influence of temperature on incidence remains a clinically relevant endpoint. The increasing CRE trend possibly reflects improved surveillance. To minimize incorrect attribution of CRE increases to predictors, we included a temporal term to adjust for this trend. The temporal resolution of antibiotic use data did not align with infection data. However, it enabled exploring antibiotic use as a predictor. Consistent with prior findings [7], our analysis with various meropenem use time lags showed meropenem use was positively associated with CRE infections six months later. Analyses adjusting for hospitalization, mobility, infection origin, and bacterial species were limited by data availability [2]. In South Korea, temperature, humidity, and mobility often peak together during warmer months, suggesting temperature may influence seasonal behavior and not necessarily directly modulate CRE. The observed 9.4% increase per 10°C rise should be interpreted with caution, as it reflects a statistical association, not a direct biological mechanism.

Despite these constraints, our findings support growing evidence that environmental factors influence AMR infections [8]. Incorporating climate variables in surveillance may improve burden estimates and guide public health interventions. Further research is warranted to identify mechanisms where temperature influences AMR.

## Conclusion

Our national surveillance-based study demonstrates that minimum ambient temperature was associated with CRE incidence, independent of antibiotic use. While based on data from South Korea, these findings add to the growing global evidence that climate factors may shape antimicrobial resistance patterns. Integrating environmental determinants such as temperature into AMR surveillance could strengthen preparedness and prevention strategies not only in South Korea but also in Europe and worldwide.

## Data Availability

All data used in this study are publicly available. CRE (Carbapenem-Resistant Enterobacteriaceae) data were obtained from the Infectious Disease Portal of the Korea Disease Control and Prevention Agency (https://dportal.kdca.go.kr/pot/is/inftnsdsEDW.do). Population data were retrieved from the Korean Statistical Information Service (KOSIS) of Statistics Korea (https://kosis.kr). Meropenem usage data were sourced from the 2022 annual report of 58 participating hospitals in the Korea National Antimicrobial Use Analysis System (KONAS; https://www.konas.or.kr:44538/xe/report). Land surface temperature data were obtained from the Terra Moderate Resolution Imaging Spectroradiometer (MODIS) Land Surface Temperature/Emissivity Daily (MOD11A1) Version 6.1 product, which provides 1-kilometer spatial resolution (https://lpdaac.usgs.gov/products/mod11a1v061/).

## References

1. Lim J, Sim J, Lee H, Hyun J, Lee S, Park S. Characteristics of Carbapenem-resistant Enterobacteriaceae (CRE) in the Republic of Korea, 2022. Public Health Wkly Rep. 2024/01/25 ed. 2024 Jan;17(4):115–27. Available from: 10.56786/PHWR.2024.17.4.1

2. Logan LK, Weinstein RA. The Epidemiology of Carbapenem-Resistant Enterobacteriaceae: The Impact and Evolution of a Global Menace. J Infect Dis. 2017 Feb 15;215(suppl_1):S28–36. Available from: 10.1093/infdis/jiw282

3. MacFadden DR, McGough SF, Fisman D, Santillana M, Brownstein JS. Antibiotic resistance increases with local temperature. Nat Clim Chang. 2018 Jun 1;8(6):510–4. Available from: 10.1038/s41558-018-0161-6

4. McGough SF, MacFadden DR, Hattab MW, Mølbak K, Santillana M. Rates of increase of antibiotic resistance and ambient temperature in Europe: a cross-national analysis of 28 countries between 2000 and 2016. Vol. 25, Euro Surveill. 2020. p. 1900414. Available from: 10.2807/1560-7917.ES.2020.25.45.1900414

5. Li W, Liu C, Ho HC, Shi L, Zeng Y, Yang X, et al. Association between antibiotic resistance and increasing ambient temperature in China: an ecological study with nationwide panel data. Lancet Reg Health West Pac. 2023 Jan 1;30. Available from: 10.1016/j.lanwpc.2022.100628

6. Kim JY, Park S, Kim EO, Chang E, Bae S, Kim MJ, et al. The seasonality of carbapenemase-producing Enterobacterales in South Korea. J Hosp Infect. 2023 Oct 1;140:87–9. Available from: 10.1016/j.jhin.2023.07.010

7. Ryu S, Klein EY, Chun BC. Temporal association between antibiotic use and resistance in Klebsiella pneumoniae at a tertiary care hospital. Antimicrob Resist Infect Control. 2018 Jul 16;7(1):83. Available from: 10.1186/s13756-018-0373-6

8. Fernández Salgueiro M, Cernuda Martínez JA, Gan RK, Arcos González P. Climate change and antibiotic resistance: A scoping review. Environ Microbiol Rep. 2024 Oct 1;16(5):e70008. Available from: 10.1111/1758-2229.70008

